# Quantifying bias from reverse causation in observational studies of dementia risk factors: A simulation study informed by age-specific reverse Mendelian Randomization

**DOI:** 10.64898/2026.02.21.26346807

**Authors:** Jingxuan Wang, Sarah F. Ackley, Ruijia Chen, Katrina L. Kezios, Adina Zeki Al Hazzouri, Deborah Blacker, Jacqueline M. Torres, M. Maria Glymour

## Abstract

**Background:** The long preclinical phase of dementia can bias estimated effects of baseline exposures on dementia incidence. We demonstrate simulations informed by reverse Mendelian randomization (MR) findings to quantify the age-specific magnitude of reverse causation bias in analyses in observational studies of the effects of body mass index (BMI) on dementia.

**Methods:** We simulated longitudinal trajectories of BMI and dementia risk from ages 45 to 90 years, calibrating to published evidence on age-specific dementia incidence, BMI, and associations of dementia genetic risk with BMI. Under the null that BMI does not influence dementia and an alternative that BMI at any age increases subsequent dementia risk, we simulated hypothetical cohort studies (n=20,000, average 15 years of follow-up), varying age of entry from 45 to 80 years. In each hypothetical cohort, the association of z-standardized BMI at study entry and dementia incidence were estimated using Cox proportional hazards models. Bias was quantified using the ratio of observed to true hazard ratios (RHRs). All scenarios were replicated 500 times.

**Results:** In the absence of a causal effect of BMI on dementia, when follow-up began at age 65 years, the RHR was 0.91 (95% CI: 0.90-0.92). When follow-up began at age 80 years, the RHR decreased to 0.68 (95% CI: 0.67-0.69), indicating substantial bias attributable to reverse causation.

**Conclusion:** Reverse causation, presumably arising from preclinical dementia, can induce substantial bias in estimates of the association between baseline exposures and dementia incidence. Simulations provide a convenient tool to quantify this bias.

## Introduction

Dementia is the seventh leading cause of death worldwide and a major contributor to disability and dependency among older adults.^1^ Given the limited effectiveness of available treatments, understanding modifiable risk factors is critical to prevent or delay the onset of dementia.^2^ Observational studies play a key role in this effort, yet they are vulnerable to reverse causation bias.^3^ Because dementia develops gradually over decades, with a long preclinical phase preceding clinical diagnosis, associations observed in late-life cohorts may partially reflect the effects of early disease processes on those exposures.^4–6^ Such bias can be superimposed on any true causal effect of the exposure on dementia incidence.

A well-known example is body mass index (BMI). Although higher mid-life BMI is linked to an increased risk of dementia, individuals with preclinical dementia often experience unintentional weight loss years before diagnosis.^6,7^ Consequently, late-life observational studies often find that higher BMI appears protective,^8–11^ but this may reflect reverse causation: weight loss due to preclinical neurodegenerative changes and associated changes in cognition and mood.^12^ If reverse causation is substantial, it may obscure a true effect of BMI at older ages on subsequent dementia risk, leading to inappropriate advice to older adults about how to best reduce their dementia risk. The 2024 Lancet Commission review of dementia risk factors for example, identified mid-life obesity as a risk factor for dementia but not obesity in old age.^2^ Quantifying bias from reverse causation in such settings would potentially allow us to identify the true causal effect of exposures at any age on subsequent dementia incidence.

Reverse Mendelian randomization (MR) uses genetic variants established to influence the outcome of interest, such as dementia, as instrumental variables to test whether the outcome causally influences potential risk factors.^13^ In dementia research, reverse MR has been used to evaluate bidirectional relationships between Alzheimer’s disease or dementia genetic liability and traits such as BMI, social isolation, and hearing impairment, providing evidence that some late-life associations may arise from reverse causation rather than causal effects of the exposure.^14–17^ However, while these studies suggest the presence of reverse causation bias, none have quantified its magnitude.

Therefore, we used simulated data informed by previously published reverse MR studies to quantify bias arising from reverse causation in observational studies of dementia risk factors, varying age when the risk factor was measured. We focused on BMI as an illustrative exposure and examined how the magnitude of bias varied across scenarios defined by age when BMI was measured and the presence or absence of a true causal relationship between exposure and outcome. This simulation-based approach provides insight into the potential magnitude of reverse causation bias and its implications for interpreting associations reported in observational dementia research.

## Methods

### The simulated cohort study

We simulated a hypothetical cohort study of BMI on dementia incidence. The cohort comprised 5,000 participants aged 45 years. We generated longitudinal trajectories of the exposure and outcome from ages 45 to 90 years, according to the data generating rules. For each simulation, we chose a baseline age for study entry for all participants and measured participants’ BMI at baseline. We followed each participant from baseline for a random period of time, drawn from a uniform distribution between 1 and 29 years, with an average follow-up of 15 years. Age was used as the time scale, and dementia incidence was generated annually.

### Causal scenarios

We investigated two causal scenarios (Figure 1). Directed acyclic graphs (DAGs) depict the assumed causal relationships in each simulation scenario. In both scenarios, the genetic risk score for dementia influences each participant’s latent preclinical dementia score, which is operationalized as a continuous score. Latent preclinical dementia score is also influenced by confounders. Confounders included age, sex, education, one measured variable C, and one unmeasured variable U. The latent preclinical dementia score in turn influences BMI trajectory for each participant, and dementia genetic risk does not affect BMI other than through latent preclinical dementia score. BMI is also influenced by both confounders C and U. In the “no causal relationship” scenario, BMI does not affect dementia incidence. In the other scenario, BMI increases the dementia incidence with a constant effect on annual incidence probability regardless of age when BMI is measured.

**Figure 1.**
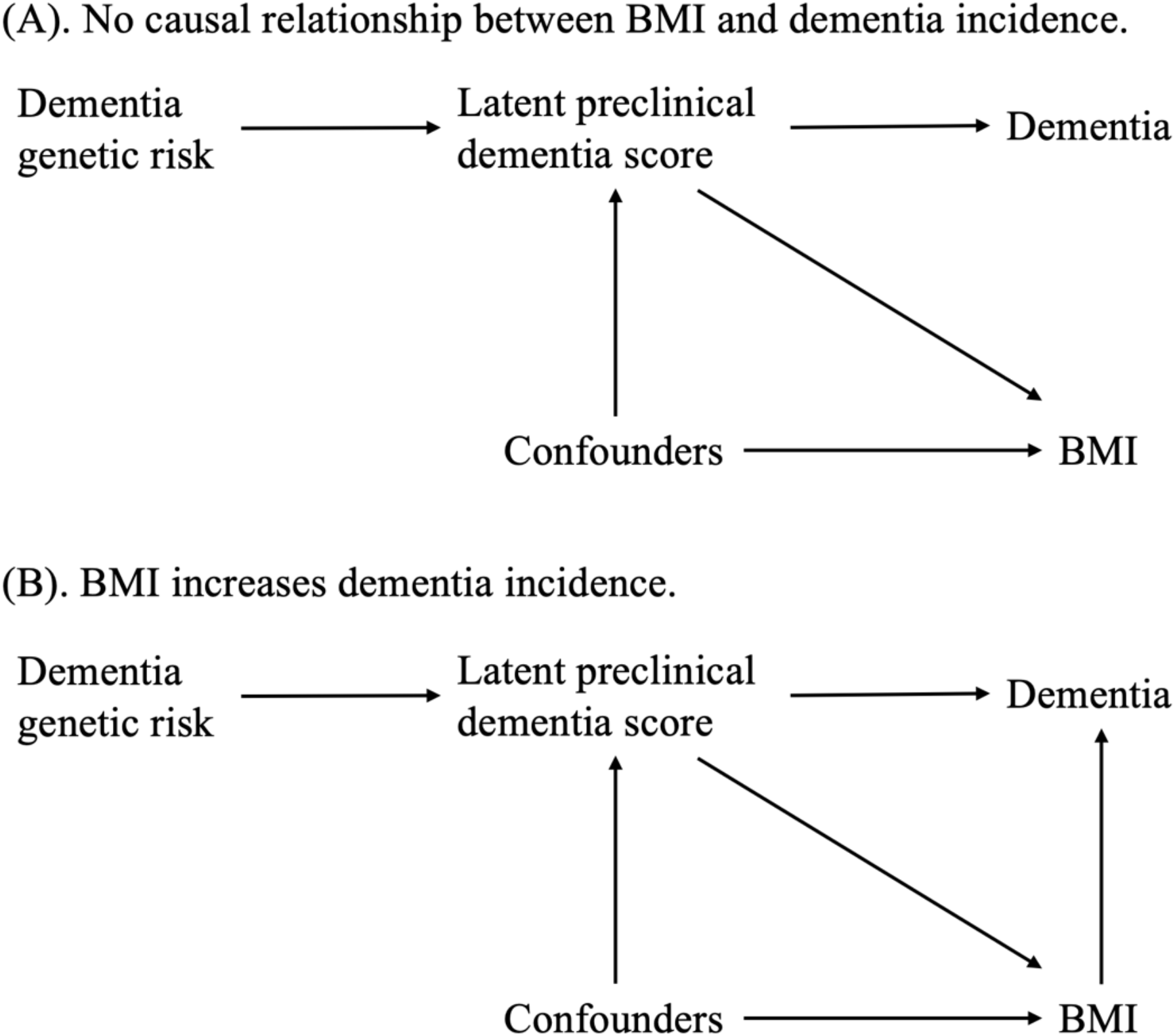
Causal scenarios under investigation.

### Data generating process

Data generating rules were calibrated to align with previously published evidence on age-specific dementia incidence, BMI trajectories across age, and age-specific associations of dementia genetic risk with BMI from reverse MR studies.^14,18,19^ We assigned each participant values of the following variables drawing a random value from a plausible distribution: a dementia genetic risk score from a normal distribution N(0,1); sex (0=male, 1=female) from a Bernoulli distribution Bernoulli(0.5); education level (0=less than high school, 1=high school or above) from a Bernoulli distribution Bernoulli(0.4); a measured confounder C from a normal distribution N(0,3); and an unmeasured confounder U from a normal distribution N(0,1). Latent preclinical dementia score was generated from dementia genetic risk score and confounders such that higher genetic risk score, older age, female, less education, lower C and higher U contribute to higher latent preclinical dementia score. For each participant *i* at age *t*,

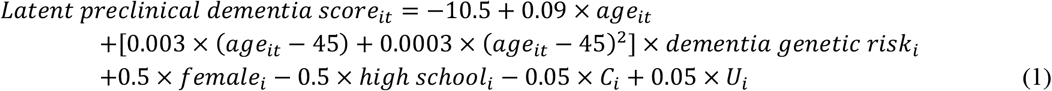

Dementia incidence was generated annually using a random Bernoulli distribution with mean probability from the latent preclinical dementia score through a logistic link

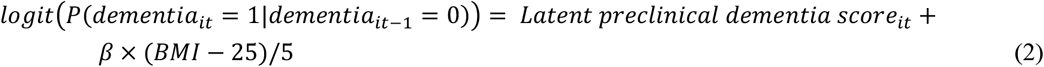

where *β* is the log hazard ratio of BMI on dementia. In the “no causal relationship” scenario, *β* = 0; in the second causal scenario, *β* = log (1.5), implying that each 5-unit increase in BMI was associated with a 5% increase in the odds of annual dementia incidence at each year of age, conditional on remaining dementia-free up to that age. The functional form of latent preclinical dementia score was selected to calibrate dementia incidence to a 2021 study that reported age-specific dementia incidence rates from clinical administrative claims in a large national Medicare population.^18^ The BMI trajectory was generated for each participant such that the inverted-U relationship between BMI and age was calibrated to a 2021 study that reported BMI trajectories from a pooled analysis of four national population-based longitudinal cohort studies.^19^ The associations between dementia genetic risk and BMI were calibrated to a 2021 reverse MR study that reported age-specific dementia genetic risk-BMI associations.^14^

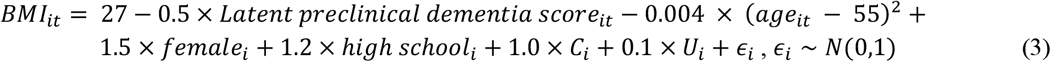

In both scenarios, we assumed no mortality or loss to follow-up until the end of study.

### Statistical analysis

In each scenario, we generated the longitudinal trajectories of BMI, latent preclinical dementia score, and dementia incidence from 45 to 90 years for each participant. We censored participants at the incidence of dementia or at the end of study when they reached age 90. To investigate the change in the magnitude of bias depending on baseline age when BMI was measured, we varied the baseline age at the start of follow-up from 45 to 80 years, with 5-year intervals. We excluded participants who had dementia incidence before each study baseline. We z-standardized BMI at study entry. The associations between BMI and dementia incidence were estimated using Cox proportional-hazard models. In the no causal relationship scenario, we considered binary obesity status in addition to continuous BMI by dichotomizing the BMI at 30 (no obesity if BMI <30 and obesity if BMI ≥30). We fit Cox models to estimate the associations between obesity and dementia incidence.

For each scenario, we repeated the simulation 500 times. To quantify the magnitude of bias caused by reverse causation, we compared the observed hazard ratio with the true effect of BMI on dementia incidence (as specified in the data generation rules) in simulated data and calculated the ratio of hazard ratios (RHR). Uncertainty in the mean bias was quantified using propagation of uncertainty based on the law of total variance, summing the within-replicate estimation variance and between-replicate Monte Carlo variability, consistent with established principles for the design and reporting of simulation studies.^20^

## Results

Across simulation replicates, the generated data reproduced age-specific patterns of BMI trajectories and dementia incidence consistent with empirical studies (Supplementary Figure 1). BMI followed an inverted U-shaped trajectory with age, declining for example from a mean of 31.0 (SD = 3.23) at age 45 to a mean of 27.4 (SD = 3.24) at age 90. Dementia incidence in the simulated data increased sharply at older ages, with an annual incidence rate of 2.8 per 1,000 person-year at age 50 to 43.8 per 1,000 person-year at age 80. The associations between dementia genetic risk and BMI increased with age, reflecting the increasing influence of latent preclinical disease on BMI trajectories cumulatively at later ages.

Figure 2 shows simulation results for the ratio of observed to true hazard ratios (RHR) across baseline age under causal scenarios with and without a causal effect of BMI on dementia. Corresponding estimated hazard ratios (HRs) are shown in Supplementary Figure 2. Under the scenario in which BMI does not influence dementia risk, the RHR was close to 1.0 (no bias) at baseline ages of 45-55. The RHR decreased with increasing age at baseline, falling to 0.91 (95% CI: 0.90-0.92) when BMI was measured at age 65 and 0.68 (95% CI: 0.67-0.69) when BMI was measured at age 80.

**Figure 2.**
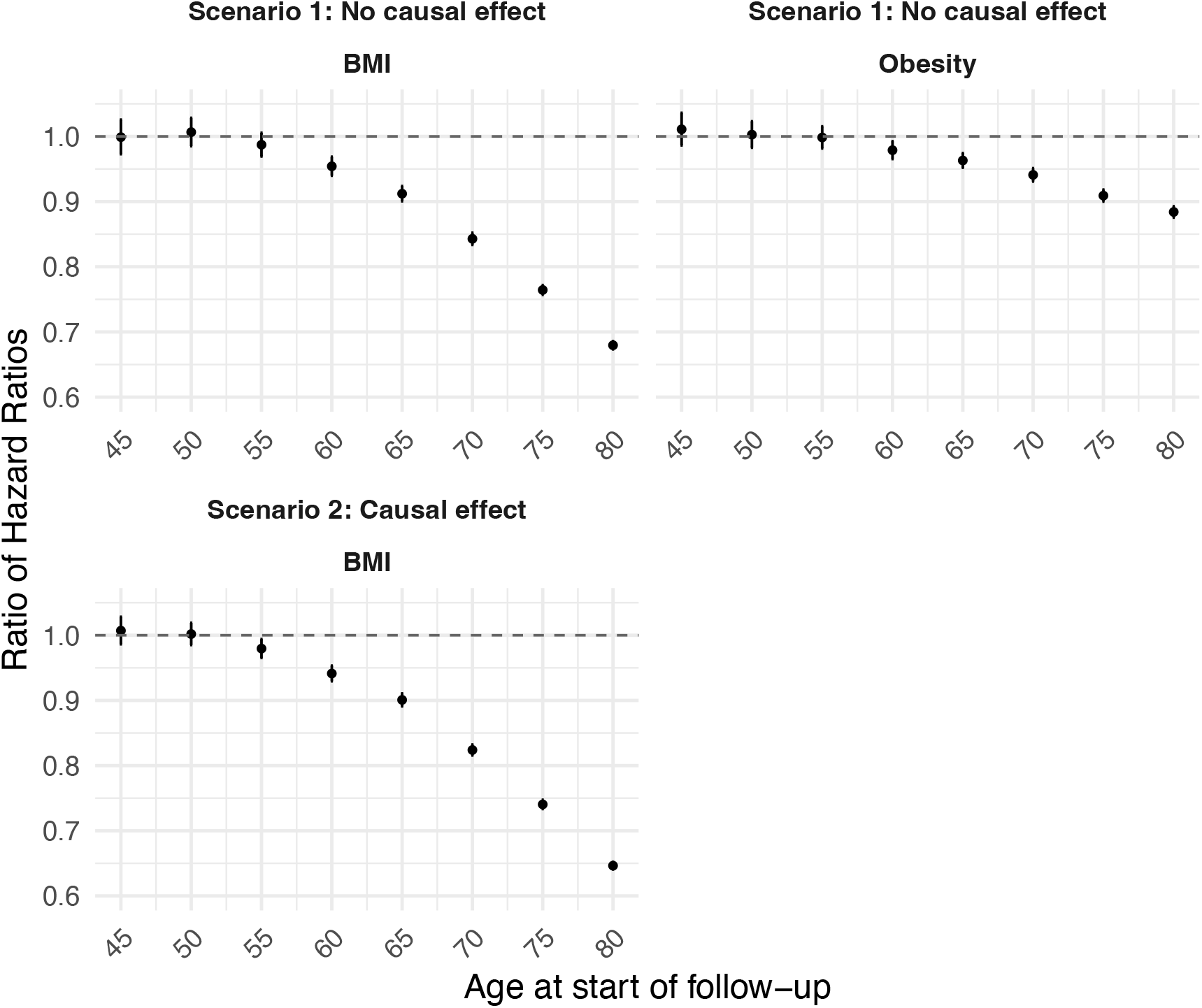
Simulation results for the ratio of observed to true hazard ratios.

A similar but less pronounced age-related decline was observed when obesity was the exposure. RHR estimates remained closer to 1.0 across most ages compared with BMI, despite a decreasing trend similar to that for BMI. At age 80, RHR reached 0.88 (95% CI: 0.88-0.89) for obesity, suggesting a modest degree of bias even in the absence of a true causal effect, but to a lesser extent than observed for BMI.

When a true causal effect of BMI on dementia incidence was encoded in the simulations (HR = 1.5 per 5-unit increase in BMI), the pattern was similar: Estimated associations were slightly biased toward the null at younger baseline ages and this bias grew with older baseline ages; the estimated effect even fell below the null -- implying that higher BMI is associated with lower dementia risk -- at older baseline ages (Figure 2; Supplementary Figure 2). At baseline age 80, the RHR was 0.65 (95% CI: 0.64-0.65).

## Discussion

We used simulations informed by prior reverse Mendelian randomization studies to quantify the magnitude of bias due to reverse causation in estimating the association between BMI and obesity with dementia incidence. Across two causal scenarios, regardless of whether there was a true causal effect of BMI on dementia incidence, we found that reverse causation arising from preclinical dementia induced substantial bias in estimated associations. The magnitude of bias varied systematically by baseline age at study entry and by exposure definition, with limited bias observed at younger baseline ages and larger bias observed at older baseline ages. For example, when study entry occurred at age 80 years, the ratio of hazard ratios deviated substantially from the null, with a value of 0.68 (95% CI: 0.67-0.69) in the “no causal relationship” scenario. In the simulated data-generating process, latent preclinical dementia influenced BMI trajectories years before diagnosis, leading to spurious associations depending on the timing of exposure measurement. Later study entry increased the likelihood that BMI was measured during the preclinical phase, thereby leading to larger reverse causation bias.

Our results are consistent with patterns observed in the epidemiologic literature. Observational studies that measured BMI in midlife and initiated follow-up earlier in adulthood have generally reported positive associations between higher BMI and dementia risk.^9,11,21–25^ For example, one meta-analysis reported a pooled risk ratio of 1.33 for dementia among individuals with midlife (ages 35-65) BMI above the obesity threshold compared with those with normal BMI.^24^ An earlier meta-analysis similarly reported an elevated dementia risk associated with midlife overweight BMI compared with normal BMI (risk ratio = 1.26).^9^ In contrast, studies focusing on BMI measured later in life have often reported null or inverse associations, a pattern frequently attributed to reverse causation. Using 38 years of follow-up from the Framingham Heart Study Offspring Cohort, Li et al. found that higher BMI at ages 40-49 years was associated with increased dementia risk, whereas no association was observed at ages 50-59 years, and a trend toward an inverse association emerged at ages 70 years and older.^11^ Similarly, a large pooled analysis reported hazard ratios per 5 kg/m^2^ increase in BMI of 0.71 (95% CI: 0.66-0.77), 0.94 (95% CI: 0.89-0.99), and 1.16 (95% CI: 1.05-1.27) when BMI was assessed 10 years, 10-20 years, and more than 20 years before dementia diagnosis, respectively. Consistent with these empirical patterns, our simulation suggests that even in the absence of a causal effect of BMI on dementia, an inverse association may be observed when BMI is measured late in life. For example, when follow-up began at age 75 years in the no causal effect scenario, the observed hazard ratio was 0.76 (95% CI: 0.76-0.77). Taken together, these findings suggest that observational studies with BMI measured earlier in life are more likely to reflect the true causal relationship with dementia risk, whereas BMI measured at older ages is increasingly susceptible to reverse causation bias arising from preclinical disease processes.

Our findings have direct implications for interpreting observational studies of BMI or obesity and dementia. Estimates derived from cohorts with older baseline ages may be particularly vulnerable to reverse causation, even when long follow-up is available. These results in combination raise the possibility that elevated BMI even in old age might have a harmful effect on dementia risk, but we have been unable to observe this effect due to reverse causation. Rigorously evaluating this possibility is important for routine guidance to older adults, who may not consider BMI a relevant risk factor.

Although our simulations focused on BMI, the same bias mechanism may apply to other exposures influenced by preclinical dementia or associated with dementia genetic risk, suggesting broader relevance of these findings. Many risk factors for dementia appear to attenuate or even reverse at older ages, such as hypertension and physical activity.^26–28^ Although reverse causation is commonly acknowledged as a potential bias, and even used to inform analytic decisions such as burn-in-periods, to our knowledge this is the first effort to quantify reverse causation bias across ages of risk factor measurement. Careful consideration of baseline age and exposure timing is therefore critical when interpreting observed associations among older adults.

Strengths of this study include the explicit encoding of causal assumptions using directed acyclic graphs, calibration of the data-generating process to multiple empirical sources, and isolation of reverse causation as a single bias mechanism. However, several limitations should be noted. We assumed no mortality or loss to follow-up and specified a constant effect of BMI on dementia risk, which may not fully reflect real-world complexity.^29,30^ In addition, the latent preclinical dementia score is an abstract construct that cannot be directly observed in empirical data. Furthermore, although the data-generating process was informed by reverse Mendelian randomization studies, the estimated associations between dementia genetic risk and BMI in those studies may themselves be affected by selection bias or selective survival, and thus may not perfectly represent real-world mechanisms.

In conclusion, our simulations show that reverse causation arising from preclinical dementia can induce substantial bias in estimates of the association between BMI and dementia incidence, particularly when BMI is measured later in life. Beyond quantifying bias for BMI, this work demonstrates how simulation studies can be used to investigate bias mechanisms that are difficult to evaluate empirically. As with all simulation studies, the validity of our results depends on the assumed causal structures and parameterization of the data-generating process. By using BMI for this demonstration, we were able to rely on well-established data on age trajectories of BMI. Future research should explore alternative simulation scenarios, incorporate competing risks such as mortality, and extend this framework to other exposures potentially affected by preclinical dementia processes.

## Data Availability

All data produced in the present work are contained in the manuscript

