## Supplement for "Quantifying bias from reverse causation in observational studies of dementia risk factors: A simulation study informed by age-specific reverse Mendelian Randomization"

**Online supplement**

**Supplementary Figure 1. Calibration of simulated data to real-world evidence**

(A). Associations between dementia genetic risk score and BMI.

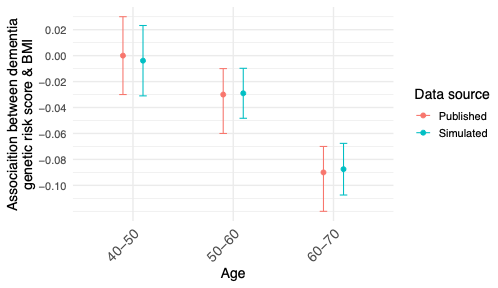

Note: Published estimates are from Brenowitz et al.^14^

(B). BMI trajectories.

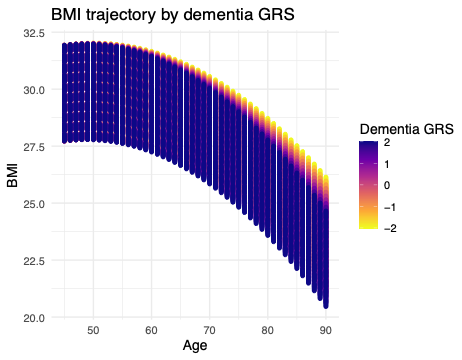

Note: BMI and age show an inverted-U relationship with peak at 55 years, as suggested by Yang et al.^19^

(C). Age-specific dementia incidence.

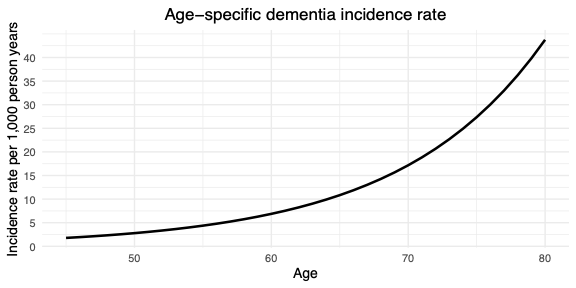

Note: Age-specific dementia incidence increases with age, with values rising gradually in midlife and accelerating rapidly at older ages, particularly after age 70, as suggested by Olfson et al.^18^

**Supplementary Figure 2. Estimated hazard ratios in the simulations**

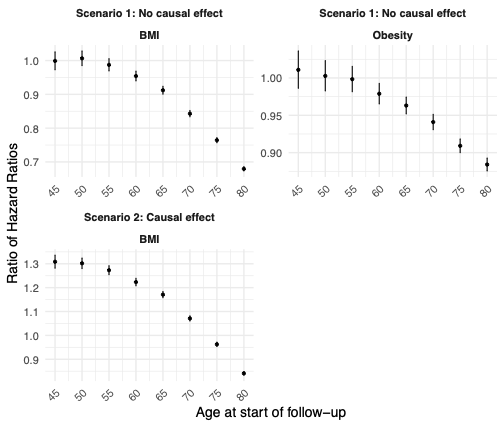
